# Post COVID-19 sequelae: A prospective observational study from Northern India

**DOI:** 10.1101/2021.06.28.21259658

**Authors:** Shivdas Naik, Manish Soneja, Soumendra Haldar, Netto George Mundadan, Prerna Garg, Ankit Mittal, Devashish Desai, Praveen Kumar Trilangi, Sayan Chakraborty, Nazneen Nahar Begam, Bisakh Bhattacharya, Ganesh Maher, Swathi S Kumar, J Kirthana, Bharathi Arunan, Ankesh Gupta, Niranjan Mahishi, Chaitra Rajanna, Prateek Parsoon, Nikhil A Kumar, Sayan Maharatna, Akashneel Bhattacharya, Vishakh C Keri, Sameer A Samed, AK Adarsh, Imtiyaz Shareef, Neeren Ravela, Satish Swain, Radhika Sarda, Harshith B Kadnur, Ashok Dudhwal, Ayush Agarwal, Kartik Vedula, Ashish Gupta, Shubham Agarwal, R Anand, Pratima Lalikar, Pallavi Jagtap, B Premjeet, Parul Kodan, Prayas Sethi, Animesh Ray, Pankaj Jorwal, Arvind Kumar, Neeraj Nischal, Sanjeev Sinha, Ashutosh Biswas, Naveet Wig

## Abstract

**Background:** Long COVID, or post-COVID-19 sequelae, is being seen in a growing number of patients reporting a constellation of symptoms, both pulmonary and extrapulmonary. Studies on COVID-19 recovered patients are scarce. Thus, there is a need to add granularity to our existing knowledge about the course and long-term effects of the infection.

**Aim:** To describe the clinical details and risk factors of post-COVID sequelae in the North Indian population.

**Method:** This prospective observational study was conducted at a tertiary healthcare centre in Northern India between October 2020 to February 2021. Patients aged >18 years with a confirmed COVID-19 disease were recruited after at least two weeks of diagnosis and interviewed for any post-COVID-19 symptoms.

**Results:** Of 1234 patients recruited, who were followed up for a median duration of 91 days (IQR: 45-181 days), 495 (40.11%) patients had symptoms. In 223 (18.1%) patients, the symptoms resolved within four weeks, 150 (12.1%) patients had symptoms till twelve weeks, and 122 (9.9%) patients had symptoms beyond twelve weeks of diagnosis of COVID-19. Most common long COVID-19 symptoms included myalgia (10.9%), fatigue (5.5%), shortness of breath (6.1%), cough (2.1%), disturbed sleep (1.4%), mood disturbances (0.48%) and anxiety (0.6%). The major determinants of developing post-COVID-19 symptoms in the patients were hypothyroidism and the severity of the disease.

**Conclusion:** Most often, patients complain of myalgias, fatigue, dyspnoea, cough and disturbed sleep. Patients who are hypothyroid or have recovered from moderate to severe COVID-19 are at higher risk of developing post-COVID sequelae. Therefore, a multidisciplinary approach is required to diagnose and manage COVID-19 recovered patients.

## Introduction

COVID-19 is a multisystem disease. The respiratory system bears the maximal brunt of the direct viral damage, but the viral infection may wreak havoc on all major systems in the body. Even after recovery from the disease, widespread respiratory, circulatory, neurological, and musculoskeletal complaints may persist. The epidemio-pathological basis of these ‘Post-COVID’ complaints is incompletely understood. These post-COVID sequelae can be due to the damage inflicted by the virus itself, by widespread damage to cytokine storm, by the immune response of the body, as a side effect of the therapy used to treat the disease, due to underlying co-morbidities or a combination of all above. The role of psychosocial factors like prolonged isolation, fear of infecting family members, socio-economic disturbances, and the stigma associated with the infection may have far-reaching consequences on mental health and well-being. (1)

As per NICE guidelines, post-COVID sequelae have been divided into acute COVID-19, ongoing symptomatic COVID-19 and the post-COVID syndrome. (2) The term ‘long COVID’ is commonly used to describe signs and symptoms that continue or develop after acute COVID-19. It includes both ongoing symptomatic COVID-19 and post-COVID-19 syndrome.

A relatively new entity, the epidemiology of post-COVID sequelae are not well known. (3) We performed this prospective observational study to describe the incidence, natural history and risk factors of post-COVID sequelae in the population of Northern India.

## Methodology

### Study design and settings

This prospective observational study was conducted in a tertiary care centre in New Delhi, India, between October 2020 and February 2021.

### Participants

The study included patients of age more than 18 years who had a confirmed COVID-19 infection (as per WHO definition) at least two weeks prior to the enrolment, with admitted patients being called after their discharge. (4) These patients were interviewed using a pre-designed proforma in the post-COVID clinic. Patients who did not visit the clinic were interviewed telephonically.

### Data collection

For teleconsultation, the contact numbers of study subjects were taken from the hospital records at the time of their diagnosis or admission for COVID-19 care. The interview was done by residents trained in administering the questionnaire and communicated with the patients in the regional language. Verbal consent was taken before the enrolment. Confidentiality of subjects was ensured throughout the study. The study protocol was approved by the institutional ethics committee.

The questionnaire included demographic details, symptoms, hospitalisation and oxygen use during acute diseases and symptomatology subsequent to recovery from acute COVID19. Patients were systematically asked about a list of post-COVID symptoms (dyspnea, myalgia, fatigue, anosmia, ageusia, chest pain, cough, mood disturbances etc.), but they were also free to report any symptoms that they considered relevant. A follow-up interview was conducted after one month and three months to look for the resolution of symptoms or any new events. Patients could visit the clinic at any time if they desired.

The severity of acute COVID-19 was defined as mild, moderate or severe as per national guidelines. (5) The post-COVID-19 symptoms of the patients were classified as per NICE guidelines. (2) Acute COVID-19 has been defined as patients with symptoms and signs of COVID-19 for up to 4 weeks. Ongoing symptomatic COVID-19 includes patients with symptoms and signs COVID-19 from 4 to 12 weeks. Patients with symptoms and signs that develop during or after an infection consistent with COVID-19, which continued for more than 12 weeks and are not explained by an alternative diagnosis are said to have Post-COVID-19 syndrome. Long COVID was defined as signs and symptoms that persist or develop after acute COVID 19 and included both “ongoing symptomatic COVID 19” and “post-COVID 19 syndrome”.

The data was analysed using STATA 16.0 software. Categorical variables were presented as N (%),are presented as mean (standard deviation [SD]) or median (interquartile range [IQR]), where ever applicable. Statistical significance was set at a p-value of <0.05. For the comparison of variables, two groups were considered. Group 1 included patients who developed the post-COVID-19 syndrome, and Group 2 included patients who did not develop the post-COVID-19 syndrome. Categorical variables were compared using the fishers exact test or chi-square test, where ever applicable, and continuous variables were compared using an independent sample Student’s t-test. In addition, a binary logistic regression model was developed to assess the impact of different variables on the likelihood of developing post-COVID-19 syndrome with the forward conditional method. Independent variables which had a p-value of <0.2 were included for binary logistic regression.

## Results

A total of 2243 patients were contacted, of whom 1645 patients responded. Of these, 147 were excluded as they were aged <18 years, and 264 patients/ attendees did not consent to the study. So, 1234 patients who recovered from COVID-19 were recruited. These patients were followed up for a median duration of 91 days (IQR: 45-181 days) after diagnosis of the disease. We were able to follow 276 patients (26%) for more than six months post-discharge/ post-end of isolation, 264 (24%) were followed for 3-6 months of discharge, 398 (37%) had a follow up till only three months, and only 140 (13%) patients had follow-up till less than one months of post-discharge/ post-end of isolation. More than 50% of the patients were followed more than two times in the clinic, once between 2 to 4 weeks and once at the end of 3 months post-discharge. About 32% of the patients were contacted for the first time after three months of their discharge. And remaining patients were contacted only once, up to 3 months.

Our cohort had a mean age of 41.4 ± 14.2 years and included 856 (69.4%) males. Of these 1234 patients, 1059 (85.8%) patients had mild COVID-19 disease, 135 (10.9%) had moderate, and 40 (3.3%) patients had severe COVID-19 disease. In addition, there were 352 (28.5%) patients who had at least one co-morbidity. The most common co-morbidities were diabetes mellitus (13.4%), hypertension (8.1 %) and hypothyroidism (5.8%). (Table 1).

**Table 1:**
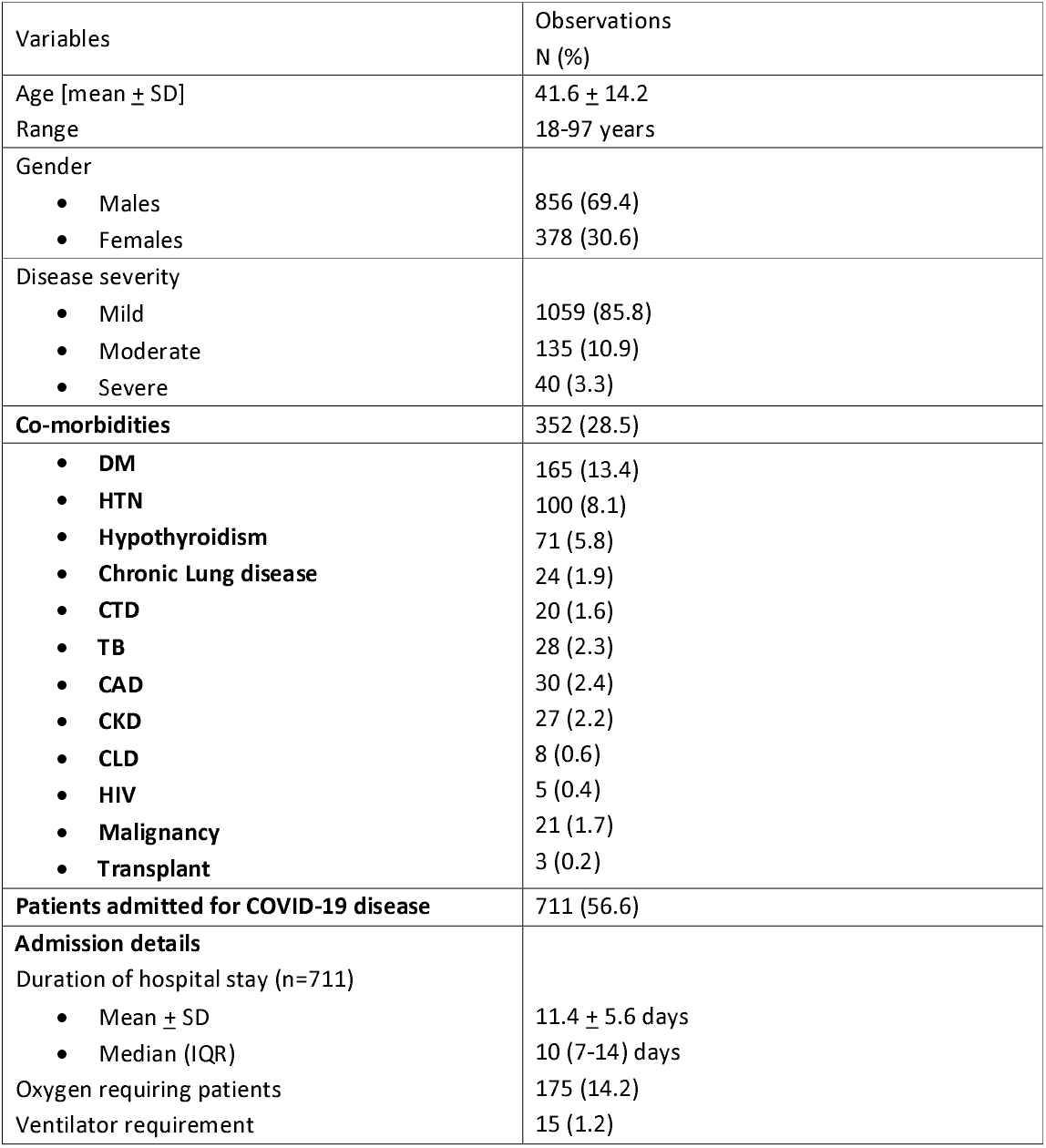
Demographic profile

A total of 711 (56.6%) patients were hospitalised for COVID-19, with a mean duration of 11.4 ± 5.6 days, with a range of 1-36 days. Oxygen support was required by 175 patients (14.2%), of whom 15 patients (1.2%) required ventilatory support. (Table-1)

Of these 1234 patients, 495 (40.11%) patients had symptoms after their discharge or end of quarantine (Fig-1, Table-1). In 223 (18.1%) patients, the symptoms resolved within four weeks. The remaining 272 (22.0%) patients had long COVID. Of all patients with long COVID-19, 150 (12.1%) patients had symptoms till twelve weeks, and 122 (9.9%) patients had symptoms beyond twelve weeks. The most common long COVID-19 symptoms included myalgia (10.9%), fatigue (5.5%), shortness of breath (6.1%) and dry Cough (2.1%). Other symptoms included disturbed sleep (1.4%), mood disturbances (0.48%) and anxiety (0.6%) (Table-2; Figure 2). The proportion of patients reporting persistent symptoms diminished with a longer time to follow-up. (Table-3)

**Figure 1:**
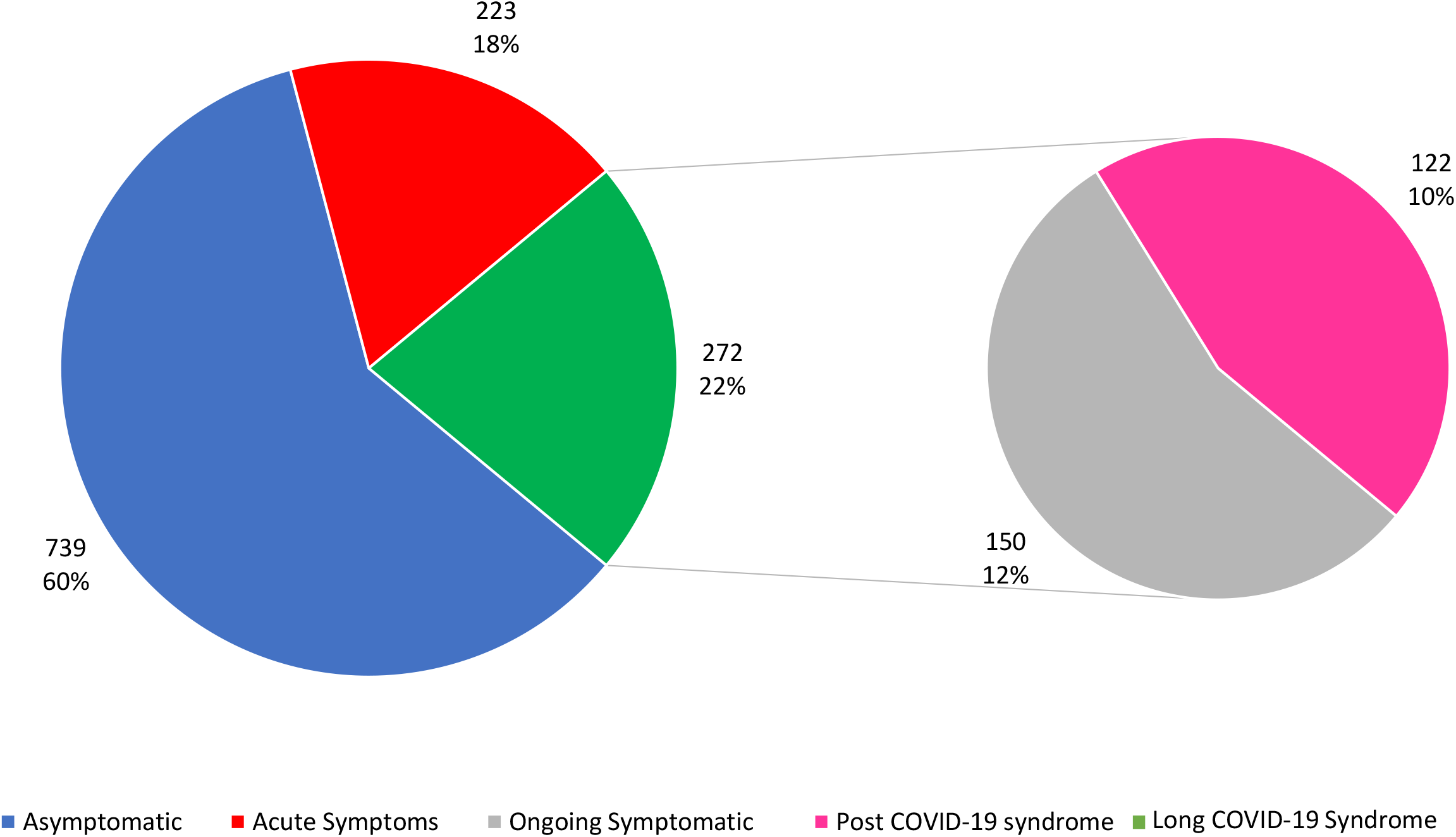
Post COVID-19 symptoms in the study group. [N=1234]. The blue portion represents patients who did not have any symptoms. The red portion represents the patients with acute COVID-19. The green portion represents patients with long COVID-19 symptoms, which included patients with ongoing symptoms, represented as the grey portion, and post COVID-19 syndrome, represented as the pink portion.

**Table 2:** Characteristics of post covid manifestations among the study sample

| Symptoms | Acute Symptoms<br>N (%) | Post COVID-19 Symptoms |  |
| --- | --- | --- | --- |
|  |  | Ongoing Symptoms<br>N (%) | Post COVID-19<br>syndrome<br>N (%) |
| Loss of Taste | 24 (1.9) | 0 | 0 |
| Loss of Smell | 16 (1.3) | 2 (0.16) | 0 |
| Anxiety | 9 (0.7) | 4(0.3) | 4(0.3) |
| Low Mood/<br>Depression | 11 (0.9) | 4 (0.32) | 2 (0.16) |
| Chest Pain | 11 (0.89) | 8 (0.7) | 6 (0.5) |
| Sleeplessness | 32 (2.6) | 13 (1.1) | 4 (0.3) |
| Cough | 34 (2.8) | 14 (1.1) | 12 (1) |
| Dyspnea | 162 (13.1) | 33 (2.7) | 42 (3.4) |
| Fatigue | 121 (9.8) | 45 (3.7) | 22 (1.8) |
| Myalgia | 145 (11.6) | 80 (6.5) | 54 (4.4) |
| Headache | 17 (1.4) | 0 | 0 |

**Figure 2:**
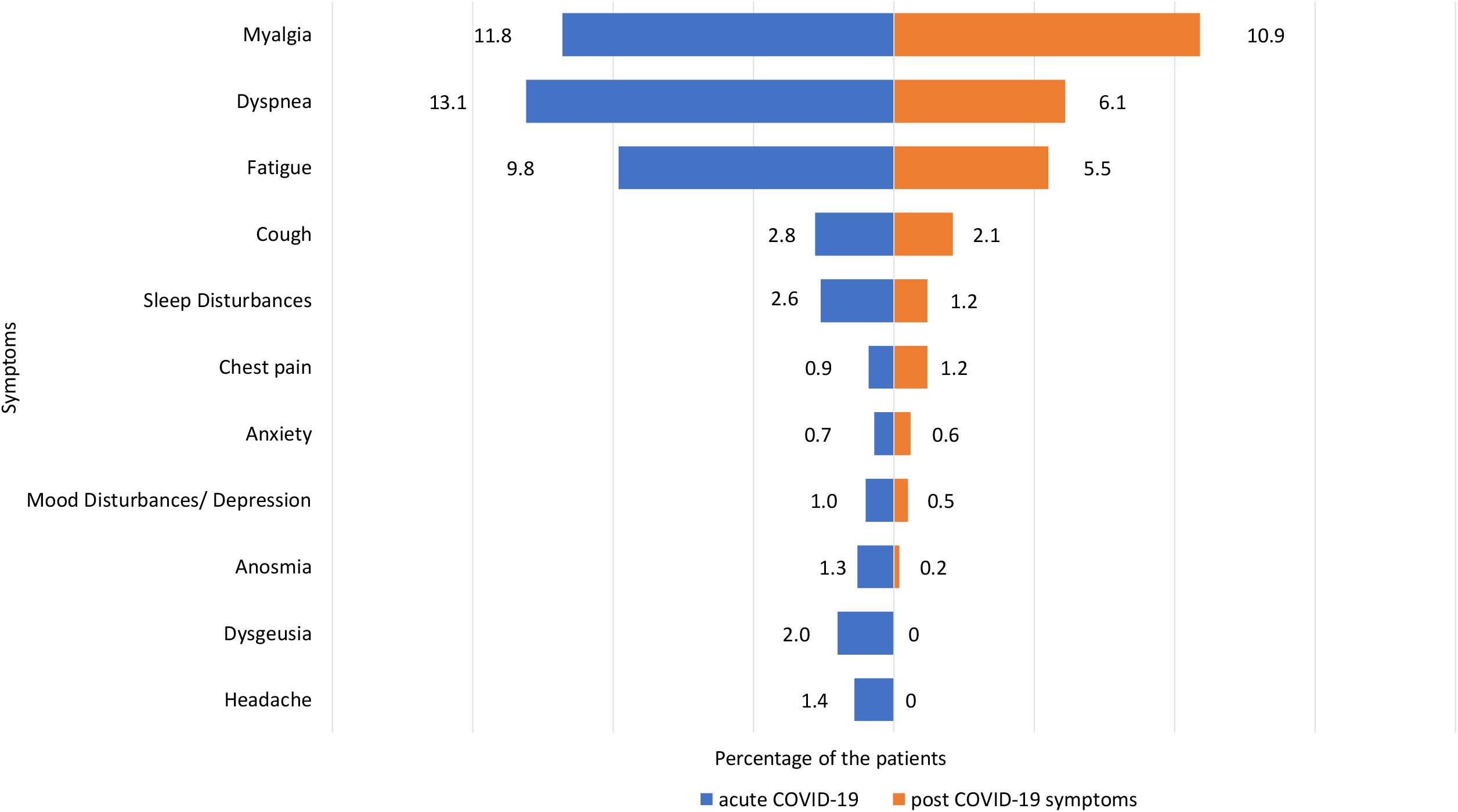
Patients (as a percentage of all patients recruited) with symptoms after discharge. The blue graph represents the patients (in percentage) with symptoms less than four weeks of diagnosis, and the red graph represents the patients (in percentage) with symptoms beyond four weeks of diagnosis.

**Table-3:**
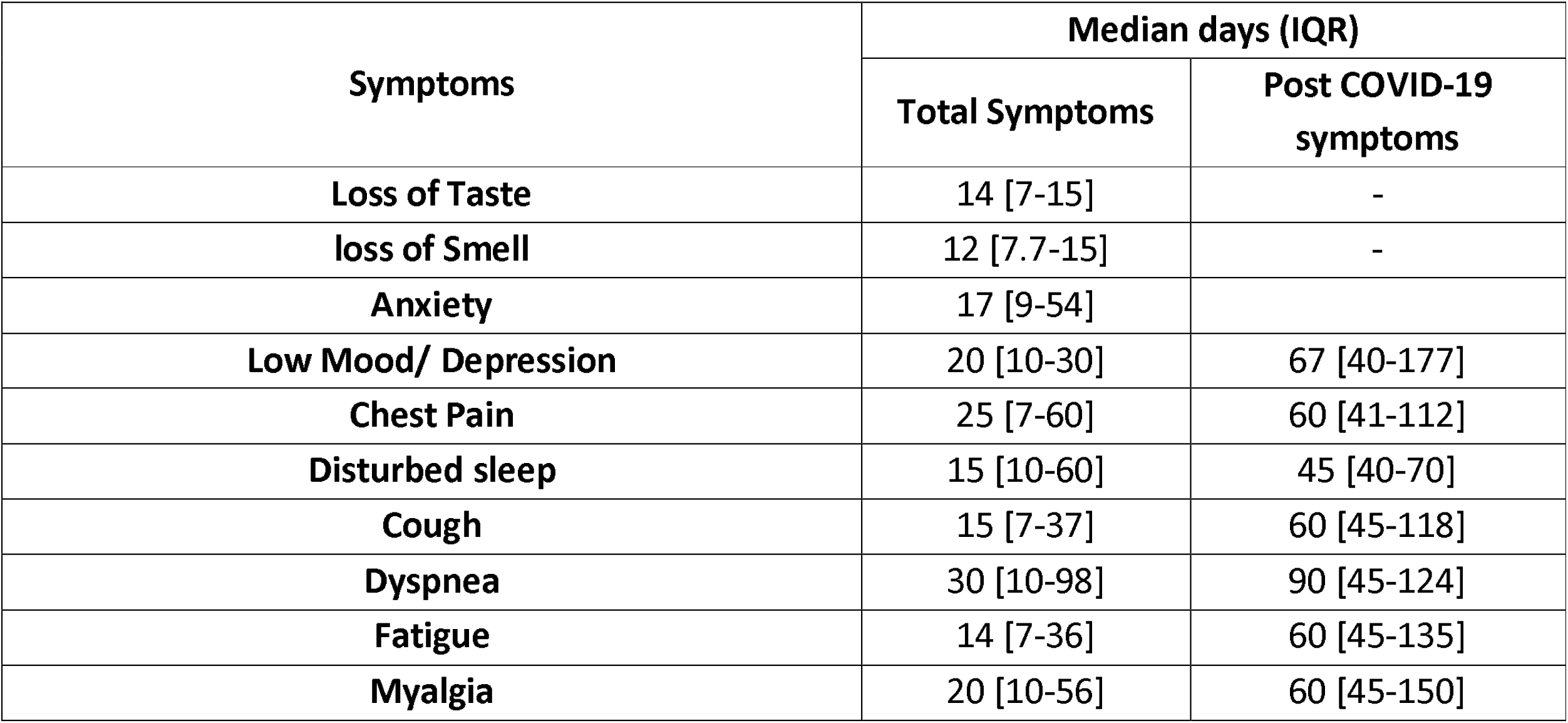
Duration of post covid symptomatology in the study group

The major determinants of developing post-COVID-19 symptoms were hypothyroidism (OR: 4.13, 95% CI: 2.2-7.6, p-value <0.001) and disease severity. Compared to mild disease, the patients who had either moderate or severe COVID-19 disease had a significantly higher chance of developing long COVID-19 symptoms (OR: 1.7, 95% CI: 1.1-2.4, p-value 0.012). Also, age was found to be tending towards significance (OR: 1.1, 95% CI: 0.99-1.02, p-value 0.056) (Table-4).

**Table-4:**
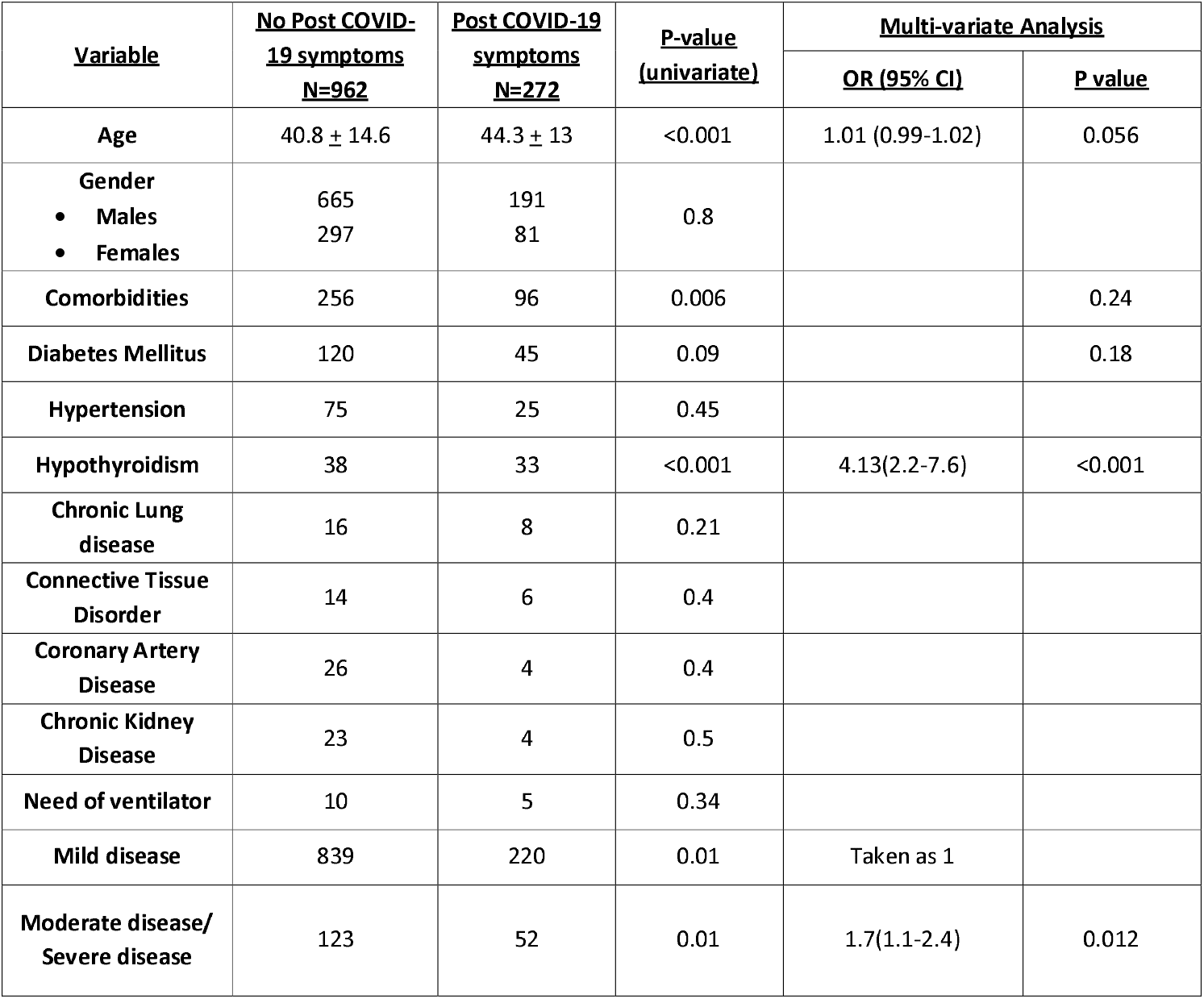
Multi-variate analysis for risk of developing COVID-19 symptoms

## Discussion

The COVID-19 pandemic is one of the greatest threats faced by humanity in recent times. The immeasurable loss of life and livelihood has left deep scars on the human psyche. Even amongst survivors, prolonged symptoms have been noted. These post-COVID symptoms significantly affect the quality of life in patients. (6) Long COVID, or post covid sequelae of COVID-19 infection, is being seen in a growing number of patients reporting a constellation of symptoms, both pulmonary and extrapulmonary, with known or undeciphered mechanisms.

In our cohort of 1234 patients, 272 (22%) patients had symptoms that persisted beyond four weeks or ‘long COVID’.(2) This is lower than previous estimates of prevalence which range from 50-80%. (7–14). Multiple reasons may have caused this; the proportions of mild and asymptomatic patients recruited in previous studies were lower; many of the studies included had a risk of selection bias, with symptomatic patients more likely to seek medical attention.

Persistent musculoskeletal complaints, including fatigue and myalgias, have been reported in 37-62% of patients symptomatic post-Covid. (2–5,7,9,10) The most common post-COVID symptoms in our cohort were myalgia (134, 10.9%) and fatigue (67, 5.5%). Studies have demonstrated frontal and cerebellar hypometabolism in patients with post-covid fatigue. (16) Decreased levels of neurotransmitters, reduced neuronal excitability, inflammation, and inhibition in the firing of motor neuron units have all been hypothesised as central factors contributing to post-covid fatigue and myalgias. (17) Metabolic factors like vitamin D deficiency, anaemia, hypothyroidism, and underlying chronic diseases also contribute to prolonged fatigue. (18) Importantly, the role of psychological influences cannot be ruled out.

The massive socio-economic upheaval caused by the pandemic has had a significant impact on the populace’s psyche. Isolation, mobility restrictions, fear of infection, financial losses, and stigma have led to mood disorders. (19,20) In our study, 49 (4%) patients reported difficulty sleeping while 17 (1.4%) patients each had depression and anxiety that occurred after recovery from the infection. However, these complaints resolved within three weeks in most patients.

Apart from psychosomatic complaints, many patients suffered from prolonged respiratory complaints, including shortness of breath (75, 6.1%) and dry cough (26, 2.1%). This is lesser than the previously estimated prevalence of 24 – 40% and 11-14% of dyspnoea and cough, respectively.(7,9,11,15) In addition to the cases discussed above, a higher number of patients with underlying respiratory conditions may have contributed to the higher prevalence in previous studies. Dyspnoea was also the most persistent complaint, with a median duration of 30 days in our cohort.

Pulmonary parenchymal injury and ARDS are hallmarks of acute COVID-19. Direct viral injury, cell and cytokine and cell-mediated injury, activation of profibrotic pathways, and trauma due to positive pressure ventilation can cause permanent scarring of lung parenchyma. (21) Follow up studies have shown that in up to 98% patients, abnormalities like ground-glass opacities, crazy paving patterns and bands of fibrosis persist in chest imaging even after 28 days from symptom onset. (22) Additionally, COVID induced thromboembolic microangiopathy, and the resulting immune-inflammatory cascade causes sizable damage to the pulmonary vasculature. (23) This may explain the high prevalence of chest pain in patients with dyspnoea post-COVID.

Neurotropism by the SARS-CoV-2 virus, neuronal inflammation mediated by direct invasion and bystander injury by the immune response to the virus, and neuroimmunomodulation through the Vagus nerve has been implicated in cough hypersensitivity causing persistent cough post-COVID. (24) This is similar to the neurogenic sensitisation that has been proposed for chronic fatigue syndrome. Thus, this may explain the co-existence of other symptoms like fatigue, myalgia and neurocognitive symptoms in post-Covid patients. (25)

The strongest determinants for post COVID symptoms in our study were the severity of the COVID19 infection (mild vs severe or moderate) and hypothyroidism. More severe illness is associated with a prolonged course of the disease and more significant damage. In patients with co-morbidities like hypothyroidism, musculoskeletal and respiratory complaints are common. Additionally, non-mild COVID-19 may induce a sick euthyroid state, recovery from which may be delayed in patients with hypothyroidism. (26) Thyroid disorder may adversely affect the disease outcome in several ways like the effect of the virus on the tissue distribution of ACE 2 receptor, increased burden of cardiovascular and psychiatric co-morbidities in turn affecting the metabolic stress.. (27) Hypothyroidism (with or without levothyroxine supplementation) have shown to negatively affect the long term outcomes in covid survivors in our study, and more prospective studies can throw more light on this association.

Another probable determinant for post-COVID symptoms was advanced age. With advancing age, decreased growth factors, stem cell migration, and cell proliferation have been demonstrated. (28) Psychosocial factors like increased isolation, poor nutrition, diminished mobility, and increased vulnerability are also more common amongst the elderly. The risk of more severe disease increases with advancing age, and our study shows that this population is also more likely to experience prolonged symptoms after the acute illness. Our study showed that probably age tended towards being a significant determinant for long COVID symptoms. Probably a higher sample size could give a better idea.

Our study is the largest prospective study of its kind, recruiting 1234 patients of all severity of Covid-19. It is also the first study to recruit a large number of patients with an asymptomatic and mild infection and thus may provide a more accurate estimate of the prevalence of post-Covid symptoms in a real-life scenario. Furthermore, since patients were actively asked about various symptoms at pre-defined intervals, recall bias was reduced. To our knowledge, our study is the first to show that hypothyroid patients are at higher risk of developing post-COVID symptoms. This highlights the need for a multispecialty approach to the diagnosis and treatment of post-COVID symptoms.

Our study had a few limitations. Loss to follow up is inherent in all cohort studies, and this was exacerbated in our study due to significant social upheaval and mobility restrictions during the pandemic. Due to mobility restrictions and limited services during the pandemic surge, data on laboratory workup and imaging of patients could not be collected systematically, which may have shed light on their pathogenesis, clinical course and management.

## Conclusion

Post COVID-19 symptoms are common and significantly distressing to patients. Most often, patients complain of myalgias, fatigue, mood disorders, and disturbed sleep. In addition, dyspnoea and cough are common and may persist for a long time. Patients who are hypothyroid or have recovered from severe COVID-19 are at higher risk of developing these symptoms. Therefore, a multidisciplinary approach is required to diagnose and manage patients with post-COVID sequelae.

## Data Availability

the data is available with the first author and can be produced on a reasonable request

## Acknowledgement

We thank all the Nursing Staffs at the Post-COVID-19 clinic, AIIMS New Delhi, for their help and support in the study.

## References

1. COVID Live Update: 174,244,202 Cases and 3,748,225 Deaths from the Coronavirus - Worldometer [Internet]. [cited 2021 Jun 8]. Available from: https://www.worldometers.info/coronavirus/?fbclid=IwAR35ZFiRZJ8tyBCwazX2N-k7yJjZOLDQiZSA_MsJAfdK74s8f2a_Dgx4iVk

2. Overview | COVID-19 rapid guideline: managing the long-term effects of COVID-19 | Guidance | NICE [Internet]. NICE; [cited 2021 Jun 8]. Available from: https://www.nice.org.uk/guidance/ng188

3. Cares-Marambio K, Montenegro-Jiménez Y, Torres-Castro R, Vera-Uribe R, Torralba Y, Alsina-Restoy X, et al. Prevalence of potential respiratory symptoms in survivors of hospital admission after coronavirus disease 2019 (COVID-19): A systematic review and meta-analysis. Chron Respir Dis. 2021 Jan 1;18:147997312110022.

4. Laboratory testing for 2019 novel coronavirus (2019-nCoV) in suspected human cases [Internet]. [cited 2021 Jun 25]. Available from: https://www.who.int/publications-detail-redirect/10665-331501

5. ClinicalManagementProtocolforCOVID19dated27062020.pdf [Internet]. [cited 2021 Jun 25]. Available from: https://www.mohfw.gov.in/pdf/ClinicalManagementProtocolforCOVID19dated27062020.pdf

6. Fernández-de-las-Peñas C, Palacios-Ceña D, Gómez-Mayordomo V, Cuadrado ML, Florencio LL. Defining Post-COVID Symptoms (Post-Acute COVID, Long COVID, Persistent Post-COVID): An Integrative Classification. Int J Environ Res Public Health. 2021 Mar 5;18(5):2621.

7. Cares-Marambio K, Montenegro-Jiménez Y, Torres-Castro R, Vera-Uribe R, Torralba Y, Alsina-Restoy X, et al. Prevalence of potential respiratory symptoms in survivors of hospital admission after coronavirus disease 2019 (COVID-19): A systematic review and meta-analysis. Chron Respir Dis. 2021 Mar 17;18:14799731211002240.

8. Chakraborty T, Jamal RF, Battineni G, Teja KV, Marto CM, Spagnuolo G. A Review of Prolonged Post-COVID-19 Symptoms and Their Implications on Dental Management. Int J Environ Res Public Health. 2021 May 12;18(10):5131.

9. Iqbal FM, Lam K, Sounderajah V, Clarke JM, Ashrafian H, Darzi A. Characteristics and predictors of acute and chronic post-COVID syndrome: A systematic review and meta-analysis. EClinicalMedicine. 2021 May 24;36:100899.

10. Iwu CJ, Iwu CD, Wiysonge CS. The occurrence of long COVID: a rapid review. Pan Afr Med J. 2021 Jan 20;38:65.

11. Lopez-Leon S, Wegman-Ostrosky T, Perelman C, Sepulveda R, Rebolledo P, Cuapio A, et al. More Than 50 Long-Term Effects of COVID-19: A Systematic Review and Meta-Analysis. Res Sq. 2021 Mar 1;rs.3.rs-266574.

12. Mandal S, Barnett J, Brill SE, Brown JS, Denneny EK, Hare SS, et al. ‘Long-COVID’: a cross-sectional study of persisting symptoms, biomarker and imaging abnormalities following hospitalisation for COVID-19. Thorax. 2020 Nov;thoraxjnl-2020-215818.

13. Moreno-Pérez O, Merino E, Leon-Ramirez J-M, Andres M, Ramos JM, Arenas-Jiménez J, et al. Post-acute COVID-19 syndrome. Incidence and risk factors: A Mediterranean cohort study. J Infect. 2021 Mar;82(3):378–83.

14. Yelin D, Margalit I, Yahav D, Runold M, Bruchfeld J. Long COVID-19—it’s not over until? Clin Microbiol Infect. 2021 Apr;27(4):506–8.

15. Frequency and Variety of Persistent Symptoms Among Patients With COVID-19 [Internet]. [cited 2021 Jun 22]. Available from: https://edhub.ama-assn.org/jn-learning/module/2780376

16. Delorme C, Paccoud O, Kas A, Hesters A, Bombois S, Shambrook P, et al. COVID-19-related encephalopathy: a case series with brain FDG-positron-emission tomography/computed tomography findings. Eur J Neurol. 2020 Dec;27(12):2651–7.

17. Rudroff T, Fietsam AC, Deters JR, Bryant AD, Kamholz J. Post-COVID-19 Fatigue: Potential Contributing Factors. Brain Sci. 2020 Dec 19;10(12):1012.

18. Garg P, Arora U, Kumar A, Malhotra A, Kumar S, Garg S, et al. Risk factors for prolonged fatigue after recovery from COVID-19. J Med Virol. 2021 Apr;93(4):1926–8.

19. Rogers JP, Chesney E, Oliver D, Pollak TA, McGuire P, Fusar-Poli P, et al. Psychiatric and neuropsychiatric presentations associated with severe coronavirus infections: a systematic review and meta-analysis with comparison to the COVID-19 pandemic. Lancet Psychiatry. 2020 Jul;7(7):611–27.

20. Brooks SK, Webster RK, Smith LE, Woodland L, Wessely S, Greenberg N, et al. The psychological impact of quarantine and how to reduce it: rapid review of the evidence. Lancet. 2020 Mar 14;395(10227):912–20.

21. McDonald LT. Healing after COVID-19: are survivors at risk for pulmonary fibrosis? Am J Physiol Lung Cell Mol Physiol. 2021 Feb 1;320(2):L257–65.

22. Shaw B, Daskareh M, Gholamrezanezhad A. The lingering manifestations of COVID-19 during and after convalescence: update on long-term pulmonary consequences of coronavirus disease 2019 (COVID-19). Radiol Med. 2020 Oct 1;1–7.

23. Dhawan RT, Gopalan D, Howard L, Vicente A, Park M, Manalan K, et al. Beyond the clot: perfusion imaging of the pulmonary vasculature after COVID-19. Lancet Respir Med. 2021 Jan;9(1):107–16.

24. Song W-J, Hui CKM, Hull JH, Birring SS, McGarvey L, Mazzone SB, et al. Confronting COVID-19-associated cough and the post-COVID syndrome: role of viral neurotropism, neuroinflammation, and neuroimmune responses. Lancet Respir Med. 2021 May;9(5):533–44.

25. Fukuda K, Straus SE, Hickie I, Sharpe MC, Dobbins JG, Komaroff A. The chronic fatigue syndrome: a comprehensive approach to its definition and study. International Chronic Fatigue Syndrome Study Group. Ann Intern Med. 1994 Dec 15;121(12):953–9.

26. Scappaticcio L, Pitoia F, Esposito K, Piccardo A, Trimboli P. Impact of COVID-19 on the thyroid gland: an update. Rev Endocr Metab Disord. 2020 Nov 25;1–13.

27. Thomas H Brix, Laszlo hegedus, Jesper Hallas, Lars C Lund. Risk and course of SARS-CoV-2 infection in patients treated for hypothyroidism and hyperthyroidism.THE LANCET Diabetes & Endocrinology, Feb 19, 2021.

28. Gosain A, DiPietro LA. Aging and wound healing. World J Surg. 2004 Mar;28(3):321–6.

